# Natural ventilation, low CO_2_ and air filtration are associated with reduced indoor air respiratory pathogens

**DOI:** 10.1101/2022.09.23.22280263

**Authors:** Joren Raymenants, Caspar Geenen, Lore Budts, Jonathan Thibaut, Marijn Thijssen, Hannelore De Mulder, Sarah Gorissen, Bastiaan Craessaerts, Lies Laenen, Kurt Beuselinck, Sien Ombelet, Els Keyaerts, Emmanuel André

## Abstract

Currently, the real-life impact of indoor climate, human behavior, ventilation and air filtration on respiratory pathogen detection and concentration are poorly understood. This hinders the interpretability of bioaerosol quantification in indoor air to surveil respiratory pathogens and transmission risk. We tested 341 indoor air samples from 21 community settings for 29 respiratory pathogens using qPCR. On average, 3.9 pathogens were positive per sample and 85.3% of samples tested positive for at least one. The number of detected pathogens and their respective concentrations varied significantly by pathogen, month, and age group in generalized linear (mixed) models and generalized estimating equations. High CO_2_ and low natural ventilation were independent risk factors for detection. CO_2_ concentration and air filtration were independently associated with their concentration. Occupancy, sampling time, mask wearing, vocalization, temperature, humidity and mechanical ventilation were not significant. Our results support the importance of ventilation and air filtration to reduce transmission.

## Introduction

Many pathogens causing acute respiratory infections are at least partly airborne transmitted^1–7^. Airborne transmission is almost exclusively an indoor phenomenon^5,8–10^. Its risk to susceptible attendants in a particular environment depends on aerosol production and removal, room volume and airflow patterns^8,10^.

Aerosol generation depends on the number of attendants, their respiratory activity, mask wearing, nasopharyngeal pathogen carriage and individual tendency to generate aerosols^7,11–14^. Aerosol settling time, removal and inactivation can all vary with pathogen, temperature, humidity, UV radiation, ventilation, and air filtration^8,10,15^.

There is some evidence supporting the use of ventilation to reduce infectious disease incidence. High CO_2_ concentration, which reflects poor ventilation, was directly associated with school absence due to illness^16^ and with common cold symptoms^14^. Low air exchange rates per person through mechanical ventilation were associated with higher incidence of pneumococcal disease during a prison outbreak and with a higher risk of tuberculin conversion in healthcare workers^7,17^. The evidence to support transmission reduction by means of portable air filters, which are more affordable than classical HVAC systems^18^, is more limited. They were associated with a reduced incidence of invasive aspergillosis and reduced surface contamination with *Methicillin-resistant Staphylococcus Aureus*^19,20^.

The quantification of respiratory pathogen and other bioaerosols in indoor air has been used to study the influence of environmental factors on disease transmission. This approach has the advantage of not requiring clinical follow-up of attendants. In such studies, indoor CO_2_ concentration was associated with higher detection of rhinovirus bioaerosols in ambient air^6^, higher concentration of bacterial cell wall components and culturable bacterial colony forming units^21^. The presence of an advanced mechanical ventilation system (which uses HEPA filtration, directional flow or increased air changes per hour) correlated with lower fungal colony forming units per unit volume in hospital settings, while bacterial bioaerosol loads were similar across areas with mechanical, advanced mechanical and natural ventilation in this study^22^. A more recent study reported that SARS-CoV-2 viral copies were more abundant in aerosols collected in closer proximity to an infected individual placed in a controlled environment. They also correlated positively with nasopharyngeal viral copies and ambient CO_2_. On the other hand, they correlated inversely with ventilation, air filtration and increased humidity^23^. As for air filtration, portable filters were shown to speed up the clearance of airborne particles^24,25^. Two small studies also suggested a reduction in detection of SARS-CoV-2 in ambient air but the effect was not significant^26,27^, while *Conway-Morris et al (2022)* did see a significant reduction in the detection of SARS-CoV-2 and other respiratory pathogens^28^. No study has thus far controlled for other important variables when assessing the influence of either ventilation or portable air filters on the load of respiratory pathogen bioaerosols in real-life settings.

In addition to quantifying transmission risk, sampling and testing of indoor air for respiratory pathogen bioaerosols may become an important add-on to other monitoring practices such as clinical samples, sentinel surveillance and sewage monitoring^29,30^. QPCR on ambient air has long demonstrated its ability to detect pathogen presence, concentration, viability, and genotype^5,30–33^. A recent study demonstrated the scalability of multiplex qPCR on indoor air samples from community settings to track the presence of SARS-CoV-2 and other respiratory pathogens^30^. Before this approach can be rolled out at scale, the factors influencing pathogen detection and concentration need better characterization.

We aimed to empirically identify the host, pathogen, behavioral and environmental factors which correlate with a higher respiratory pathogen bioaerosol load in indoor ambient air. We hypothesized that factors assumed to contribute to airborne transmission would be associated with higher bio-aerosol loads. If so, this would validate the use of qPCR on air samples as a proxy to quantify transmission risk and the effect of transmission reduction efforts. Also, these same factors would need consideration when performing qPCR on indoor air samples for epidemiological surveillance.

In a prospective cohort study, we therefore tested indoor ambient air from community settings for 29 respiratory pathogens using qPCR over a 7 month period. We investigated which of the following factors influenced pathogen detection and concentration: the number of attendees, attendee density (number of attendees divided by room volume), sampling duration, mask wearing, vocalization (voice use), natural ventilation (opening of doors and windows), air filtration, presence of mechanical ventilation, COVID-19 incidence, indoor CO_2_ concentration, temperature and relative humidity. In an interventional sub-study, we evaluated the effect of mobile air filters in a nursery. See *Supplementary Methods* for definitions of each assessed variable.

## Results

### Pathogen detection varies with season and age of attendants

We collected 341 environmental air samples in 21 sampling sites between October 2021 and April 2022. See *Supplementary Table 1* for sampling site characteristics. Sampling times (mean of 133 min and median of 126 min) corresponded well with the 120 min target. Two samples had missing results of the respiratory pathogen panel, while 36 had a missing result of the TaqPath SARS-CoV-2 assay (Thermo Fisher Scientific). The number of missing values for all variables is listed in *Supplementary Table 3*. Methods for inferring them are described in S*upplementary Methods*.

When comparing positivity rates of all samples, the most frequently detected pathogens, in descending order, were *Streptococcus pneumoniae* (58%), human enterovirus (incl. rhinovirus) (54%), human bocavirus (45%), human adenovirus (40%) and human cytomegalovirus (38%). The percentage of samples which were positive for at least one pathogen was highest in the 3-6 year old age group (30/30, 100%) followed by 0-3 years (122/123, 99%), 25-65 years (9/10, 90%), 12-18 years (19/24, 79%), 18-25 years (44/57, 77%), 6-12 years (21/29, 72%) and over 65 years (46/68, 68%). *Supplementary Figure 2* shows a detailed picture of the detected pathogens by age group and time-period. *Supplementary Table 1* shows that positivity rates varied significantly by sampling location, including within one age category.

Temporal variations in the positivity rates of pathogens are apparent in *Figure 1*. Human bocavirus, human cytomegalovirus, human enterovirus (incl. rhinovirus) and *Streptococcus pneumoniae* were almost always positive in the nursery setting shown. We observed a long peak of human adenovirus and *Pneumocystis jirovecii* over the winter. Other pathogens had shorter peaks, such as *Human coronavirus 229E, Human coronavirus HKU-1, Human coronavirus OC43*, enterovirus D68, influenza A virus, human parainfluenza virus 3, respiratory syncytial virus and SARS-CoV-2. For SARS-CoV-2, enterovirus D68 and influenza A virus, variations in positivity corresponded with results from clinical samples in University Hospitals Leuven, which is located adjacent to the sampling location.

**Figure 1.**
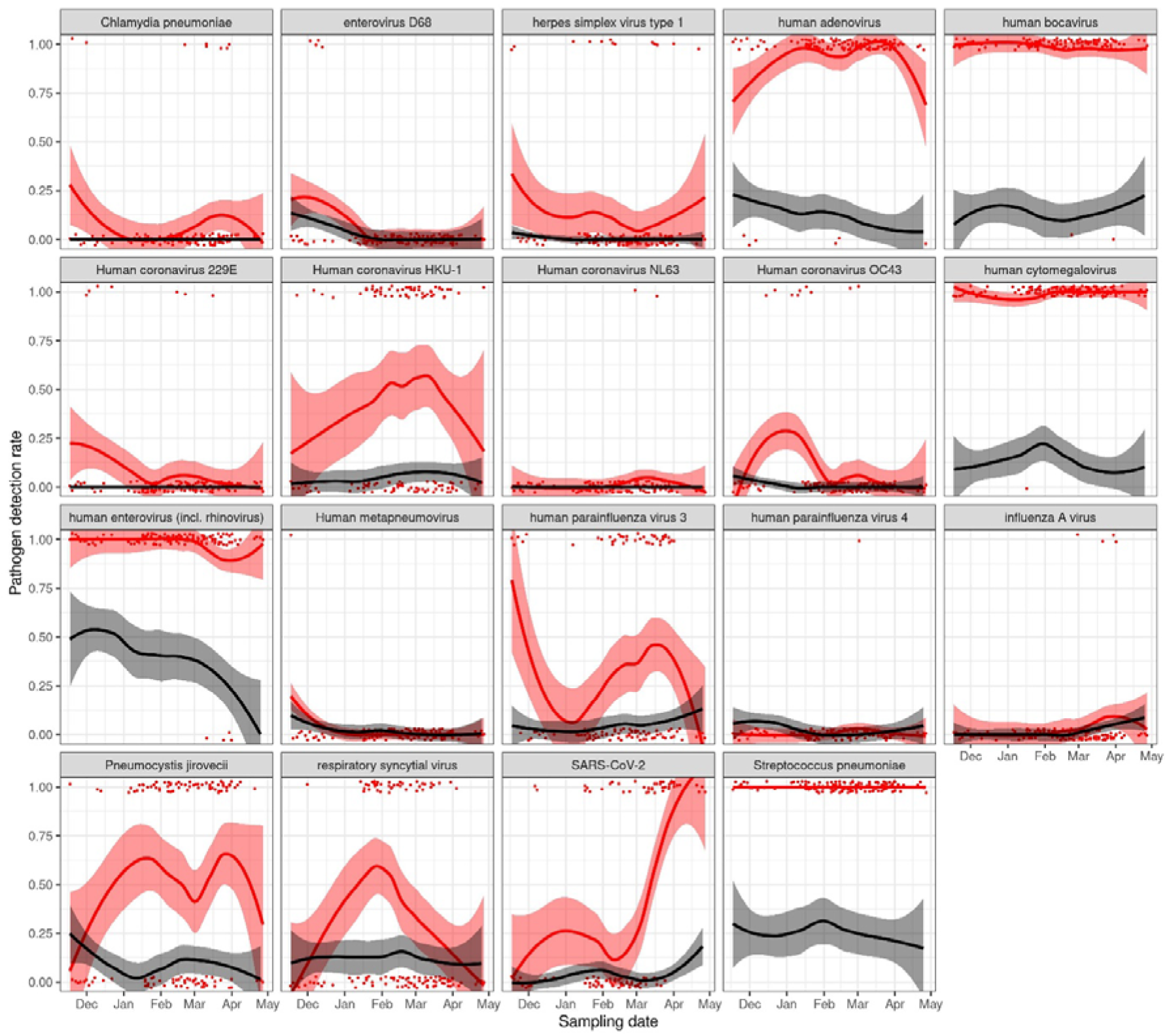
Positivity rates of respiratory pathogens in ambient air in nursery locations compared to clinical samples from a local laboratory. Each panel shows a LOESS regression of the positivity rate for each pathogen with 95% confidence intervals. Pathogens which were positive in at least one air sample are shown. Results for the ambient air are shown in red, while red dots show the individual test results. We included 121 air samples. For comparison, we retrieved the results of the 206 respiratory panels for respiratory viruses performed in University Hospitals Leuven in children between 0 and 3 years old from October 2021 to May 2022. Their results, with corresponding LOESS regression, are shown in black. The nursery was the most stable of sampling sites regarding sampling frequency, occupancy and the specific individuals present in the sampling location (Supplementary Figure 5). Supplementary Figure 6 shows the corresponding results for all sites.

**Figure 2.**
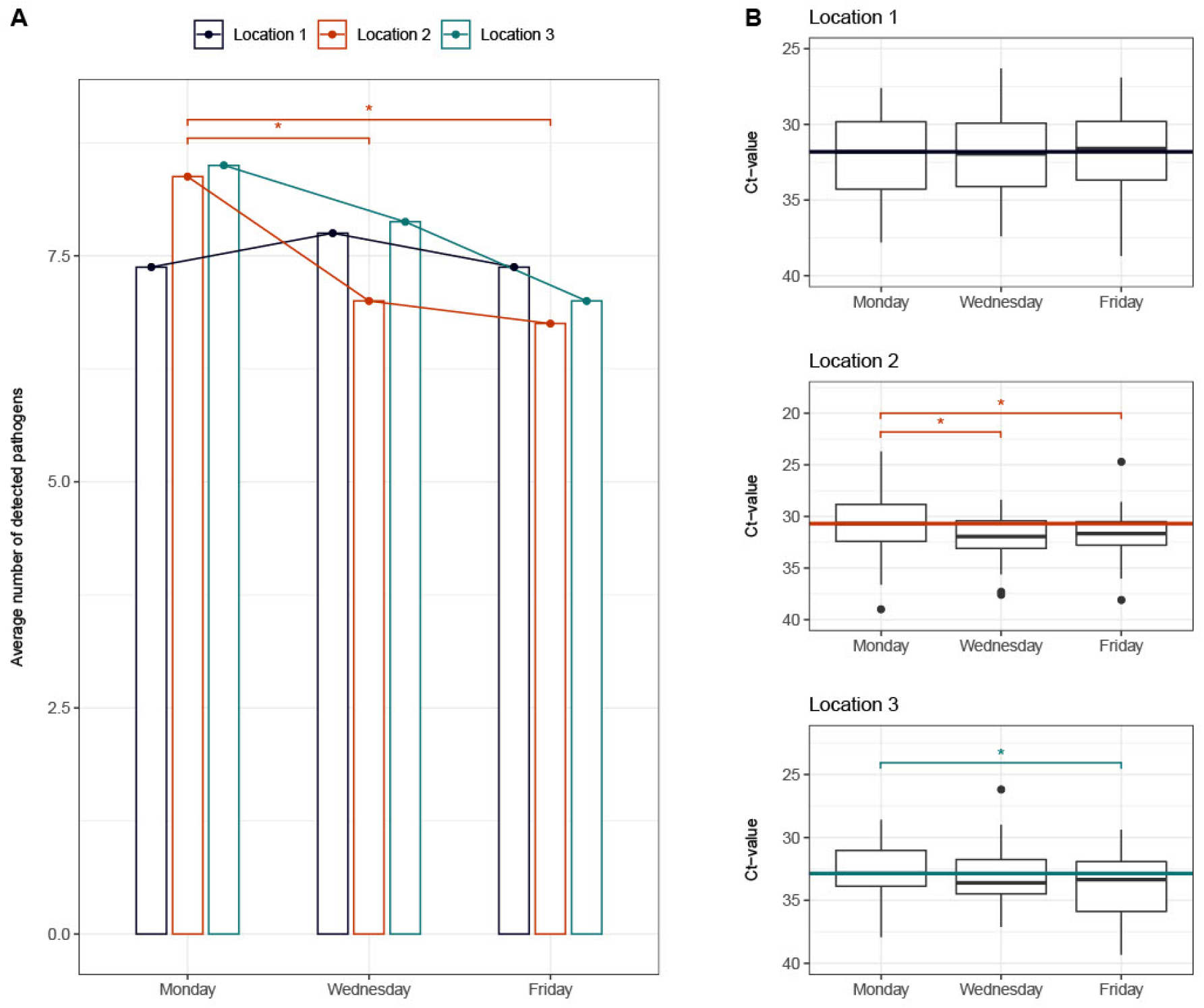
The influence of air filtration on respiratory pathogen concentration in ambient air. Location 1 = control group, Location 2 = filtration with Blue PURE 221 filters (Blueair®), Location 3 = filtration with Philips 3000i filters (Philips®). Panel a) shows the mean number pathogens detected in each of the nursery locations throughout filtration phases (Mondays = no filtration, Wednesdays = 48h of continuous filtration, Fridays = 92 hours of continuous filtration). Significant differences in a Cochran Q test are highlighted*. Panel b) shows the evolution of Ct values of all pathogens throughout filtration phases in each of the nursery locations. The horizontal line corresponds to the mean Ct value on Mondays. Significant differences in a mixed-model linear regression are highlighted*.

### The influence of host, pathogen, behavioral and environmental factors on indoor bioaerosol load

We determined independent effects of a range of variables on airborne pathogen detection and concentration, by considering the qPCR result of each pathogen in a sample as a separate observation. Pathogen type was considered a covariate in the resulting models, which included corrections for within-sample correlation. Missing data was imputed, and for each model backward elimination was performed until only statistically significant variables remained. Subsequently, observations with imputed variables were removed to confirm the observed associations.

We excluded pathogens with less than 10 positive tests after grouping them – to increase statistical power – as follows: human parainfluenza virus 1 to 4 under ‘parainfluenza viruses’; *Human coronaviruses 229E, HKU-1, NL63* and *OC43* under ‘other coronaviruses’. At least 10 positive results were present for 14 pathogens before grouping and for 12 pathogens after.

#### Factors associated with pathogen presence

First, positivity for any respiratory pathogen was the binary outcome in a logistic regression model (LRM). Backward elimination on the data including imputed datapoints left pathogen, month, age group, natural ventilation, CO_2_ and vocalization as significant variables. Contradictorily, increased vocalization was associated with decreased pathogen detection. After exclusion of observations with imputed variables from the resulting model, vocalization was removed as significant variable (*Table 1 panel a*). The odds of detecting a respiratory pathogen increased by 8.8% per 100 ppm increase in CO_2_ concentration. In contrast, odds decreased by 11% per stepwise increase in natural ventilation. Significance levels and effect sizes were similar in the mixed logistic regression model (MLR) and generalized estimating equations (GEE) models correcting for within-sample correlation (*Supplementary Table 5*).

**Table 1.** lists the pathogen, host, behavioral and environmental parameters significantly associated with indoor air bioaerosol load after backward elimination in logistic regression models (LRM). Panel a) lists the factors associated with pathogen detection, p-values and effect sizes of environmental factors (odds ratio and 95% CI). Panel b) lists the factors associated with pathogen concentration (measured in qPCR Ct values), p-values and effect sizes of environmental factors (change in Ct value and 95% CI).

| **a) Pathogen detection (all pathogens)** |  |  |
| --- | --- | --- |
| **Remaining variables** | **p-value** | **Odds ratio and 95% CI** |
| Pathogen | <0.0001 |  |
| Age group | <0.0001 |  |
| Month | 0.0025 |  |
| CO<sub>2</sub> | 0.0015 | 1.09 (CI 1.03 to 1.15) per 100 ppm increase in CO<sub>2</sub> |
| Natural ventilation | 0.0097 | 0.89 (CI 0.80 to 0.97) per step increase |
| **b) Pathogen concentration (qPCR Ct of positive samples, all pathogens)** |  |  |
| **Remaining variables** | **p-value** | **Coefficient and 95% CI** |
| Pathogen | <0.0001 |  |
| Age group | <0.0001 |  |
| Month | 0.002 |  |
| CO<sub>2</sub> | <0.0001 | -0.08 (CI -0.125 to -0.04) per increase of 100 ppm |
| Air filtration | 0.00054 | 0.58 (CI 0.25 to 0.91) |

**Table 2.** The influence of the qPCR panel on SARS-CoV-2 detection. This table shows the results of the SARS-CoV-2 qPCR targets contained in the TaqPath assay (ORF1ab, N and S) and the SARS-CoV-2 qPCR contained in the in-house multiplex qPCR for respiratory pathogens (targeting ORF1ab).

|  |  | Taqpath SARS-CoV-2 qPCR |  |  |  |  |  |  |  |
| --- | --- | --- | --- | --- | --- | --- | --- | --- | --- |
|  |  | ORF1ab |  | N gene |  | S gene |  | Any gene |  |
|  |  | Pos | Neg | Pos | Neg | Pos | Neg | Pos | Neg |
| Respiratory panel (ORF1ab) | Pos | 7 | 2 | 7 | 2 | 4 | 5 | 7 | 2 |
|  | Neg | 70 | 224 | 78 | 216 | 23 | 271 | 83 | 211 |

To assess whether these associations held true for different types of pathogens, we used the retained independent variables from these models to run a LRM with backward elimination for each pathogen. These models had less power due to lower sample sizes, however a significant association between mean CO_2_ and detection of human enterovirus (incl. rhinovirus), other coronaviruses, *Pneumocystis jiroveci* and *Streptococcus pneumoniae* remained present. Contradictorily, we found a negative association with the detection of human bocavirus. As for natural ventilation, it was negatively associated with the detection of *Pneumocystis jiroveci* and respiratory syncytial virus. *Supplementary Table 6* lists all model outcomes. *Supplementary Figure 7* shows the univariate correlations between CO_2_ or natural ventilation and pathogen detection.

#### Factors associated with pathogen concentration

Backward elimination on the data including imputed datapoints left pathogen, month, age group, CO_2_ and air filtration as significant variables. Each increase in CO_2_ by 100ppm decreased the qPCR Ct value by 0.13. Natural ventilation was not significantly associated with concentration, which contrasts with the previous analysis. On the contrary, air filtration was significantly associated with pathogen concentration, with a 0.57 increase in average Ct in its presence. Significance levels and effect sizes were similar when excluding imputed values, or when running a MLRM (*Supplementary Table 5 panel b)*.

We then ran a LRM with backward elimination for each pathogen, again taking qPCR Ct values as numeric outcome. Mean CO_2_ remained positively associated with a higher concentration (lower Ct value) of human adenovirus, human bocavirus, human cytomegalovirus, and *Streptococcus pneumoniae*. Contradictorily, it was associated with a lower concentration (higher Ct value) of respiratory syncytial virus. Air filtration was associated with lower concentrations of human bocavirus, human cytomegalovirus, other coronaviruses and *Streptococcus pneumoniae* (See *Supplementary Table 7* for all model outcomes).

#### Air filtration reduced pathogen detection and concentration in an interventional comparison

Starting from February 7th, air samples were taken simultaneously in the three nursery groups for 24 days (Mondays, Wednesdays and Fridays). Location 1 had no air filtration, location 2 had three Blue PURE 221 filters installed, with a total theoretical clean air delivery rate of 1770 m^3^/hour and a resulting number of air changes per hour (ACH) of 10.7. Location 3 had three Philips 3000i filters installed, with a total theoretical clean air delivery rate of 999 m^3^/hour and a resulting number of air changes per hour of 6.1 (*Supplementary Figure 2* and *Supplementary Table 2*).

First, we compared the positivity for any respiratory pathogen between three phases of air filtration in each group separately: no ongoing filtration (Mondays), 48 hours of continuous filtration (Wednesdays) and 92 hours of continuous filtration (Fridays). The Cochran’s Q test showed no significant difference between Mondays, Wednesdays and Fridays in location 1 (p = 0.6762). In location 2, a significant difference was present across days (p = 0.0006). Pairwise comparisons demonstrated a difference between Mondays and Wednesdays (p = 0.022900) and Mondays and Fridays (p = 0.000933) but not between Wednesdays and Fridays (p = 1). In location 3, the difference between days did not reach significance, but there was a trend (p = 0.07005).

Next, we used linear mixed-effects regression models to evaluate the change in average concentration of respiratory pathogens on Wednesdays and Fridays, compared to baseline on Mondays, in each location separately. We saw no significant change in average Ct values throughout filtration phases in location 1 (p = 0.95 when comparing Mondays to Wednesdays and 0.71 when comparing Mondays to Fridays). In location 2, there was a significant increase in Ct value of 1.22 (95% CI 0.65 - 1.79, p < 0.0001) between Mondays and Wednesdays. It remained significant when comparing Mondays to Fridays, with an increase in Ct value of 1.34 (95% CI 0.56 - 1.70, p = 0.00018). In location 3, the difference in Ct value was not significant when comparing Mondays and Wednesdays (Ct +0.33, 95% CI -0.32 - 0.98, p =0.31). However, there was a significant increase on Fridays compared to Mondays (Ct +1.02, 95% CI 0.37-1.67, p =0.003). *Supplementary table 8* lists all model outcomes.

### The qPCR platform has a major influence on pathogen detection

We compared positivity for SARS-CoV-2 with a McNemar test in 303 air samples tested with both the respiratory panel (targeting ORF1ab) and the TaqPath SARS-CoV-2 assay (targeting ORF1ab, S and N). P-values were <0.0001 when comparing the respiratory panel either to the ORF1ab target or to all targets in the Taqpath SARS-CoV-2 qPCR, revealing a major influence of the qPCR platform on the ability to detect this pathogen in indoor air.

## Discussion

In this prospective cohort study, we tested 341 indoor ambient air samples from 21 community settings for 29 respiratory pathogens using qPCR. We investigated which pathogen, host, behavioral, and environmental variables influenced their detection and concentration. In an interventional sub-study, we evaluated the effect of mobile air filters in a nursery.

### Multiplex qPCR on indoor ambient air can support surveillance of respiratory pathogens

Testing ambient air for bioaerosols of significance to human health has a long history in research^3,6,21,32^. While outdoor air sampling to detect respiratory pathogens is possible^26^, indoor air may be the more attractive alternative as humans spend over 90% of their time indoors, and as most respiratory transmission occurs there^5,8,9,34^. *Dinoi et al (2022)* combined data from 73 studies to show that the SARS-CoV-2 bioaerosol load is lowest in outdoor air, and higher in indoor air from hospitals than from community settings^31^.

During the COVID-19 pandemic, pathogen detection in sewage was scaled and provided important policy insights^29^. As environmental samples are not influenced by clinical test indications, tendency for testing or laboratory capacity, they can complement diagnostic tests and sentinel surveillance. Sewage sampling can surveil the population of entire cities, but also has disadvantages. It is highly contaminated with environmental microorganisms, the relationship between respiratory and gastrointestinal shedding may be complex, and runoff times may be long and variable^29^. These make air sampling an interesting and complimentary alternative.

A recent study demonstrated the scalability of performing multi-pathogen qPCR on air samples from community settings to highlight pathogen presence^30^. In that study and ours, pathogen presence differed by age group, which highlights the influence of host factors such as age, nasopharyngeal carriage and immunity on bioaerosol load. Pathogens such as human adenovirus, human bocavirus and *Human coronavirus OC43*, which cause childhood illnesses, were more frequently observed in schools than locations populated by other age groups in both studies. Also, both saw periodical variations in air sample positivity for influenza A virus and SARS-CoV-2, which corresponded to a change in incidence in the same geographical area (*Figure 1*). Periodical differences in positivity rates within one age category and geographical location can be explained by variations in local epidemiology, behavior, environmental factors, or a combination. This underscores the need to characterize the sampling sites to interpret the epidemiological relevance of pathogen presence in ambient air.

### Ventilation is independently associated with reduced detection and concentration of multiple respiratory pathogens

By testing more samples and pathogens than previous studies, we were able to show for the first time that both respiratory pathogen presence and concentration were independently associated with CO_2_ concentration, after correcting for a range of variables. Natural ventilation was also independently associated with pathogen detection. Results were consistent across models (LRM, GEE and MLRM). These results suggest that bioaerosol load in indoor ambient air correlates strongly with low levels of ventilation.

Pathogen specific models were generally consistent with these results, where statistical power allowed a conclusion (*Supplementary Tables 6* and *7*). Two exceptions were human bocavirus and respiratory syncytial virus. In the former, CO_2_ correlated negatively with pathogen detection, although positively with pathogen concentration. In the latter, natural ventilation correlated negatively with detection as expected, but CO_2_ was negatively correlated with concentration. Type I errors or uncorrected confounders may explain these inconsistencies. The strength of the correlation between the CO_2_ concentration and the presence of a particular pathogen was often mirrored in the strength of the inverse correlation between natural ventilation and detection of the same pathogen *(supplementary figure* 7).

These results provide strong empirical quantitative support for the use of ventilation to reduce transmission risk, consistent with previous modelling studies^8,10^. They further substantiate the need to characterize the environment, e.g. through CO_2_ measurement, when using qPCR to detecting pathogens in ambient air for epidemiological surveillance.

### Portable air filters reduce respiratory pathogen bioaerosols in ambient air

In several multivariate models, air filtration remained independently associated with a lower concentration of respiratory pathogens in our study, even after controlling for ventilation related factors (natural ventilation, the presence of an HVAC system and CO_2_ concentration).

When analysing positivity rates in the two nursery sites with air filters, we saw a significant reduction in the number of detected pathogens during filtration in the location equipped with the highest filtration capacity (theoretical air changes per hour of 10.7). The concentration of positive pathogens was also significantly reduced. Ct values increasing by 1.34 on average (95% CI 0.56 - 1.70) between Mondays, when filtration was absent, and Fridays, after 4 days of continuous filtration. In the location equipped with less filtration capacity (theoretical air changes per hour of 6.1), we saw a trend towards a reduction in the number of detected pathogens. Here, the pathogen concentration was reduced significantly after four days of continuous filtration, but not after two. On Fridays, the Ct values increased by 1.02 on average (95% CI 0.37-1.67) compared to Mondays. We saw no difference in the control group. The observed effect and dose response relationship confirm the efficacy of air filtration to reduce the respiratory pathogen bioaerosol load, given that capacity is sufficient.

### Under-detection in bioaerosol sampling

While qPCR on respiratory samples is considered the gold standard method to rule out respiratory infection, it is currently unclear to what extent air sampling can rule out the presence of infectious pathogens in the ambient air of congregate settings. Reasons for under-detection, where pathogens are not detected despite their presence in epidemiologically relevant quantities, should be considered when using air sampling for surveillance.

We demonstrated the importance of knowing the age group of attendants at a sampling site and the accurate assessment of ventilation. Other environmental factors such as temperature and humidity, whose relationships with bioaerosol load are complex and pathogen dependent, were not significant in our models, but may require consideration in specific circumstances^35^. Behavioral factors such as mask wearing and vocalization were not retained in our models, even if they are known to have an important influence on aerosol generation^36^. This may result from a lack of power or from a confounder effect, as mask wearing may coincide with the implementation of other mitigation measures.

In addition, under-detection can result from the sampling method and duration. We did not compare air samplers but did use one with a comparably high flow rate, which is an important determinant of sensitivity^37,38^. Within the narrow range in our study, the sampling time was not independently associated with pathogen detection or concentration.

We did observe the importance of using a sensitive qPCR platform, as the positivity rate of SARS-CoV-2 via an in-house respiratory panel multiplex qPCR aimed at ORF1ab was significantly less sensitive than the TaqPath COVID-19 assay for all three gene targets separately (ORF1ab, S and N) and jointly. Possible explanations are differences in the transport medium used, variations in turn-around time or properties of the panel itself.

Lastly, the concentration of airborne pathogens is known to be greater in proximity to an infectious individual^8,39^, hence the sensitivity of an air sample will depend on proximity to the source and air mixing patterns in the room. We did not record the distance of individuals to the air sampler in the current study. This problem could be circumvented by placing multiple devices in the room, using devices with higher flow rates, or by sampling within an HVAC system. The latter may facilitate the sampling of a larger number of individuals per sample^38^.

### Limitations

Our study has several limitations. First, we did not attempt to isolate replication-competent virus or collect biological samples from attendants. This limits our ability to link risk factors with the risk of transmission directly. Second, we did not determine the exact concentrations of respiratory pathogens in ambient air as no standard curves were developed for each pathogen and qPCR platform. While this does not negate the importance of significant variables, it does influence the transferability of effect sizes in terms of changes in qPCR Ct values to other settings not using the exact same qPCR panels. Third, natural and mechanical ventilation rates and airflows were not assessed directly or modelled comprehensively. This limits our ability to determine whether the proximity of attendants to the sampling device may have influenced bioaerosol detection and concentration.

## Materials and methods

### Sample collection

Between October 2021 and April 2022, we collected air in a convenience sample of community settings in and around the city of Leuven, Belgium. Sampling sites covered different age groups: nursery (0-3y), preschool (3-6y), primary school (6-12y), secondary school (12-18y), adults (18+) and nursing homes (65+). See *Supplementary Table 1* for detailed characteristics of the sampling site and *Supplementary Table 2* for descriptions of the HVAC systems present in six sites. We focused on children and the elderly because of high incidence and morbidity from respiratory infections^21–23^. For university auditoria, rooms where high CO_2_ values were registered in the weeks prior to the start of the study were selected for inclusion.

We sampled for two hours unless site specific schedules required shorter sampling (e.g. lunch time in schools). An AerosolSense active air sampler collected air in standard AerosolSense Capture Media (Thermo Fisher Scientific, Waltham, MA) (S*upplementary Figure 1*). We measured environmental parameters such as CO_2_ and humidity either manually (Testo 435-4) or every 10 minutes using a remote climate sensor (Elsys) (S*upplementary Figure 3*). We used the former for 58 samples and the latter for 283.

See *Supplementary Methods* and *Supplementary Table 3* for detailed definitions of each assessed parameter related to climate, human behavior and the environment, and the imputation of missing values.

### Sample processing and analysis

The samples were transported to the lab on the day of collection and stored at 4°C. The median processing time was 0.92 days (range 0.26 to 14.25) for the TaqPath qPCR and 3.32 days (range 0.79 to 16.23) for the multipathogen respiratory panel.

#### Nucleic acid extraction

For SARS-CoV-2, we used the MagMAX™ Viral/Pathogen II (MVP II) Nucleic Acid Isolation Kit for automated extraction (Thermo Fisher Scientific, AM1836) on 200 μl sample input. For internal control, samples were spiked with a purified MS2 bacteriophage as per manufacturer’s instructions (Thermo Fisher Scientific, A47817). Extracted RNA was eluted from magnetic beads in 50 μl MagMAX Viral/Pathogen Elution Buffer.

For the multiplex respiratory panel, Total Nucleic Acid (TNA) extraction started from 500 µl of air sample in UTM with NucliSens extraction reagents on easyMAG or eMAG (BioMérieux, Lyon, France). We used the specific B protocol on the instrument after off-board lysis for 10 minutes and continuous shaking. A 10μL mixture of Phocine Distemper Virus^24^ and Phocine Herpesvirus-1^25^ was added to the lysed sample before extraction as RNA and DNA internal controls. The elution volume of TNA was 110 µl.

#### Detection of SARS-CoV-2 in air samples by RT-qPCR (Taqpath)

*Cuypers et al. (2022)* described the method for performing the Taqpath COVID-19 CE-IVD RT-PCR assay (Thermo Fisher Scientific, Waltham, MA, USA) on air samples^26^.

#### Detection of 29 respiratory pathogens in air samples by multiplex qPCR (respiratory panel)

An in-house respiratory panel, consisting of 12 real-time multiplex PCRs, was run in 96 well plates on QuantStudio DX (Thermo Fisher Scientific, Waltham, MA, USA). The end volume of each PCR reaction mix was 20 µL: 5 µL of TNA, 5 µL of master mix (TaqMan Fast Virus Mix, Thermo Fisher Scientific, Waltham, MA, USA) and 10 µL of primer/probe mix. The temperature profile used was as follows: 50°C for 10’ followed by 20’’ at 95°C and 45 cycles of 3” at 90°C and 30” at 60°C.

The panel detects seven non-viral pathogens (*Mycoplasma pneumoniae*, C*oxiella burnettii*, C*hlamydia pneumoniae*, C*hlamydia psittaci*, S*treptococcus pneumoniae, Legionella pneumophila* and P*neumocystis jiroveci*) and twenty-two viruses (influenza A virus, influenza B virus; human parainfluenza viruses 1 to 4; respiratory syncytial virus A/B; human enterovirus (incl. rhinovirus); enterovirus D68; herpes simplex virus type 1; herpes simplex virus type 2; *Human metapneumovirus*; human adenovirus; human bocavirus; human parechovirus; *Human coronaviruses 229E, HKU-1, NL63* and *OC43*; human cytomegalovirus; Middle-East Respiratory Syndrome (MERS) virus; severe acute respiratory syndrome viruses 1/2 (SARS-CoV-1/2) through the ORF1ab target. Since all positive results for ORF1ab were attributed to SARS-CoV-2, the panel could detect 22 viruses and 29 pathogens in practice. *Supplementary table 9* lists all targets and Ct thresholds.

The specificity was validated using External Quality Control (EQC) samples, cultures and clinical samples. We followed ISO15189:2012 requirements when performing the analysis. *Supplementary Methods* describes the method for excluding non-specific amplification.

#### Detection of 29 respiratory pathogens in human respiratory samples by multiplex qPCR

To compare whether the pathogens in air samples corresponded to pathogens found in patients, we retrieved the publicly available results of respiratory panels performed on clinical samples in the national reference center for respiratory viruses during the study period^27^. They were run on respiratory samples of inpatients and outpatients at University Hospitals Leuven.

### Portable air filters

To test the efficiency of portable air filters to reduce bioaerosol load, we placed them in two separate locations in a nursery. Another separate space was the control group. Up to 20 toddlers and 1 to 4 caregivers occupied each space on working days. Toddlers were always in the same location. *Supplementary Figure 4* shows the placement of air filters.

The study assessed two types of air filters. The Blue PURE 221 (Blueair®) is a HEPA and carbon filter-based device with a clean Air Delivery Rate (CADR) of 590 m3/hour. From January 17 onwards, three devices were present in nursery location 2. On the first 7 days of air filtration in this location, the air was filtered after sampling, and sampled again several hours later. The Philips 3000i (Philips®) is another HEPA and carbon filter-based device with a CADR of 333□m3/h. From February 7 onwards, three devices were present in location 3 (*Supplementary figure 2* and *Supplementary Tables 1* and *2*). From this moment, sampling took place concurrently in all three locations. On Mondays, air filtration started after sampling. Then, air filtration continued for the rest of the week. On Wednesday and Friday, sampling was repeated in each location for 2 hours per day. Air filtration was discontinued after sampling on Friday.

### Samples inclusion and exclusion

We included all samples tested with either the Taqpath SARS-CoV-2 qPCR or respiratory panel when describing pathogen detection. To avoid duplication, only the Taqpath SARS-CoV-2 qPCR was considered for SARS-CoV-2. The Taqpatch qPCR was not performed on 35/341 samples between January 3rd and 14^th^ due to financial constraints. The Taqpath SARS-CoV-2 result was missing in one more sample and the respiratory panel in 2 more samples due to failed transport between labs. Samples without a respiratory panel result were included. *Supplementary table 3* lists the missing host, behavioral and environmental values and measures taken in their presence.

*Supplementary Methods* describes the procedure for imputing missing behavioral or environmental variables.

We only used samples with results for both the Taqpath SARS-CoV-2 and respiratory panel qPCR to compare sensitivities for SARS-CoV-2.

### Statistical analysis

To assess the influence of host, pathogen, behavioral and environmental variables on pathogen presence, we used a logistic regression model (LRM), generalized estimating equations model (GEE) and mixed logistic regression model (MLRM). Each test for a particular pathogen was one observation, while positivity was the binary outcome. Each pathogen group had equal weight. The pathogen type was a variable in the models. Both GEE and MLRM corrected for within-sample correlation of tests.

To assess the influence of the variables mentioned above on pathogen concentration, an LRM and MLRM were used. Pathogen concentration was measured by the qPCR Ct value of a positive pathogen. Again, each test for a particular pathogen was one observation, while the pathogen was considered a covariate in the model.

After imputing missing variables, as described in *Supplementary Methods*, we used backward elimination in all models to estimate effect sizes of the most important variables. 95% confidence intervals were computed as follows: coefficient estimate +/- standard error * 1.96. We used the Wald test to estimate p-values in GEE models and the Chi squared test for LRM and MRLM models. p-values were not corrected after backward elimination. After backward elimination, we removed observations with imputed variables to confirm the results.

In an exploratory analysis, we evaluated whether the influence of variables found to be significant in the above models differed by pathogen. We ran LRM models with both pathogen detection and Ct value as outcome, for each detected pathogen separately, only using the retained significant variables from the models including all pathogens.

Lastly, we evaluated the effectiveness of air filtration by focusing on repeated samples taken in the 3 nursery locations. First, we used a Cochran’s Q test to compare pathogen detection rates between three phases of air filtration for each location separately: no ongoing filtration (Mondays), 48 hours of continuous filtration (Wednesdays) and 92 hours of continuous filtration (Fridays). Pairwise Cochran’s Q tests followed when the difference in phases was significant. We used Holm correction for multiple testing. We used linear mixed-effects regression models to evaluate the effect of different air filtration phases on pathogen concentration, including week and pathogen as random effects, for each location separately. Ct values for a particular pathogen were included only if the pathogen was detected in samples from all three filtration phases during the same week. Confidence intervals were calculated using the confint command in R, p-values were obtained using the Kenward-Roger approximation of the T-distribution (pbkrtest package in R).

We used a McNemar test to compare the sensitivity of the Taqpath SARS-CoV-2 and respiratory panel qPCR for SARS-CoV-2.

A two-sided p-value of ≤0.05 was considered significant in all analyses.

## Data Availability

All data produced in the present study are available are added in supplementary file source-file.xlsx.

## Data availability

All data relating to environmental conditions and the number, types and concentrations of pathogens detected are added in supplementary file source-file.xlsx. A data dictionary is added to the same file.

## Acknowledgments

The study was financed through internal KU Leuven funds. JR acknowledges support of the Research Foundation Flanders (FWO, grant number: 1S88721N). The manufacturers of the air sampling and filtration devices were not involved in the design, conduct, analysis, or manuscript writing. The AerosolSense active air samplers, AerosolSense Capture Media and Blue PURE 221 devices were purchased. The Philips 3000i devices were donated by the manufacturer.

## Author information

### Author contributions

The study was conceptualized by JR, EK, LB and EA. HDM, LB, JR, JT and BC collected the data. Analysis was performed by CG, JT, SO, MT and JR. JR, CG, SO, JT and EK wrote the manuscript. The other authors reviewed the text.

## Ethical declarations

### Ethical approval

The study received approval from the Ethics Committee Research UZ / KU Leuven (S66518, B3222022000873). No informed consent was required from occupants of the sampled environments. Sampling took place after the management of each institution had agreed to take part in the study.

