## supplementary information for "Natural ventilation, low CO_2_ and air filtration are associated with reduced indoor air respiratory pathogens"

### Supplementary methods

#### Detailed definition of factors collected for each sample

- Predominant age group at the sampling site: 0y-3y, 3y-6y, 6y-12y, 12y-18y, 18y-25y, 25-65y, +65y. (*supplementary table 1*).
- Month of sampling.
- Mean number of attendees, measured at the start and end of each sample.
- Attendee density: mean number of attendees divided by sampling room volume (m^3^). The number of attendees was estimated by headcount both at the start and end of sampling and averaged per sample.
- Sampling time: in minutes, manual entry per sample.
- Mask-wearing: estimated by Linkert scale (no one, almost no one, minority, majority, almost everyone, everyone) at the start and end of sampling. Averaged per sample.
- Vocalization: estimated by Linkert scale (no one talks, only teacher talks, minority talks, majority talks, everyone talks, singing) at the start and end of sampling. Averaged per sample.
- Natural ventilation: estimated by Linkert scale (no natural ventilation, 1 window open, door open, multiple windows open, door and window open) at the start and end of sampling. Averaged per sample.
- Purifier: binary (enabled, disabled), manual entry per sample.
- Mechanical ventilation: binary (absent/present), manual entry per site. See *Supplementary Tables 1* and *2*.
- Weekly COVID-19 incidence Leuven: COVID-19 incidence for the city of Leuven in the seven days until the day before sampling, per 100000 inhabitants^1^.
- Mean indoor CO_2_ concentration: numeric (parts per million/ppm). Either measured manually at the start and end of each sample and averaged or measured continuously and averaged over the total sampling duration (<11 minutes before start of sampling until <11minutes after end of sampling) *(Supplementary Figure 3)*
- Mean indoor temperature: numeric (degrees Celsius, °C). Either measured manually at the start and end of each sample and averaged or measured continuously and averaged over the total sampling duration (<11 minutes before start of sampling until <11minutes after end of sampling) *(Supplementary Figure 3)*.
- Relative humidity: numeric (%). Either measured manually at the start and end of each sample and averaged or measured continuously and averaged over the total sampling duration (<11 minutes before start of sampling until <11minutes after end of sampling) *(Supplementary Figure 3)*.

#### Imputing missing data

If a manual measurement was missing at the start or end of sampling, the corresponding other value was used as average for that sample.

Missing values for temperature were inferred through the following procedure:

- We computed the mean of measurements in the same location in the same period (7 days before to 7 days after) to infer the value. Both manual and continuous measurements were included.

When there was no registered value either before or after sampling for mask wearing, ventilation or vocalization, they were inferred as follows:

- The source notes were checked for information to fill the missing datapoint.
- We computed the mean of measurements in the same location in the same period (7 days before to 7 days after) to infer the value.
- The value was inferred from memory by the data entry team.

#### Excluding possible non-specific amplification of respiratory panel results

When a qPCR CT value of the respiratory panel was above the clinically defined threshold, we repeated the qPCR with fresh primer probe mixes. *Supplementary Table 9* lists all qPCR targets, primer/probe sequences and thresholds in the respiratory panel. For each pathogen with a possible case of non-specific amplification, the five samples with lowest CT values were reanalyzed. If there were less than five occurrences of non-specific amplification, they were all repeated. If at least 60% of samples returned positive, the initial results were retained. If less than 60% of samples returned positive, we reclassified all results of samples with possible non-specific amplification for that pathogen as negative (*Supplementary Table 3*).

### Supplementary figures

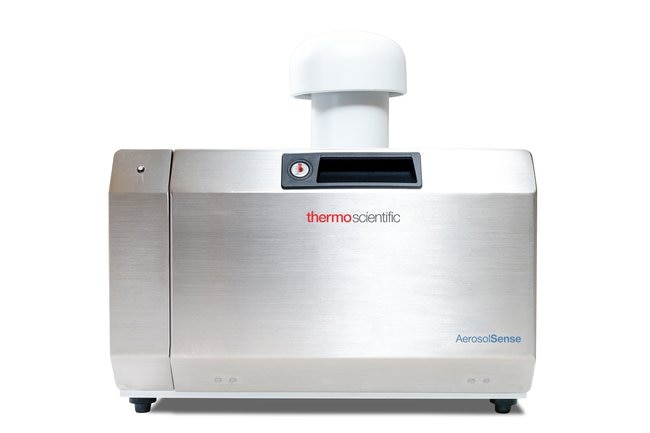

***Supplementary Figure 1. AerosolSense air capturing device (Thermofisher®).*** *This is an impaction-based active air sampler with multiple nucleic acid collection media. Air was sampled at a rate of 200 L/min through a vertical collection pipe and impacted onto the collection media. The AerosolSense Air Sampler is designed to collect aerosolized particles with a diameter between 0.1–15μm. It detected 0.32 genome copies/L of SARS-CoV-2 50% of the time during 75 minutes of sampling in a rapid deployable module with an interior volume of 28,040 L. SARS-CoV-2 was always detected at 3.2 gc/L during 8 hours of sampling in the same test room^2^.*

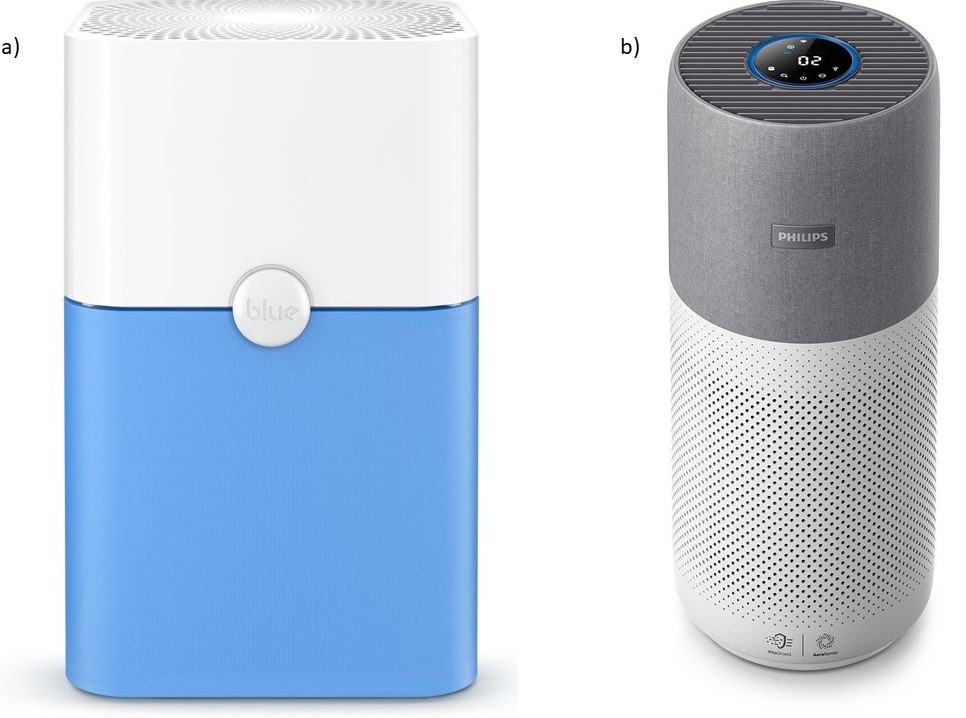

***Supplementary Figure 2. Air filtration devices.*** Panel a) shows the Blue PURE 221 (Blueair®) air cleaner, which is based on High Efficiency Particulate Air (HEPA) and carbon filtration technology*. Its Clean Air Delivery Rate (CADR) is 590 m3/hour as per manufacturer specifications.* Panel b) shows the Philips 3000i (Philips®), another HEPA and carbon filtration-based air cleaner*. Its CADR is at least 333 m3/h when operated in “turbo” mode as per manufacturer specifications. The devices* were used *in stage 2, which corresponds to a CADR of 186.7 m3/h3 per device.*

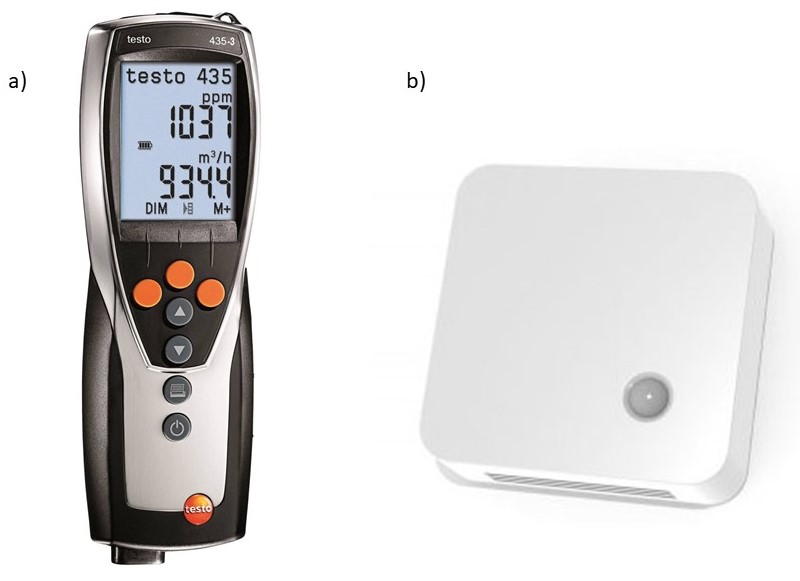

***Supplementary Figure 3. Data loggers.*** *Panel a) shows the hand-held Testo 435-4 multifunctional indoor climate meter (Testo®), which was connected to an indoor air quality probe for CO_2_, temperature, humidity and absolute pressure^4^. Panel b) shows the LoRAWAN connected monitor (Elsys®) which monitored indoor CO_2_ levels, temperature and humidity continuously^5^.*

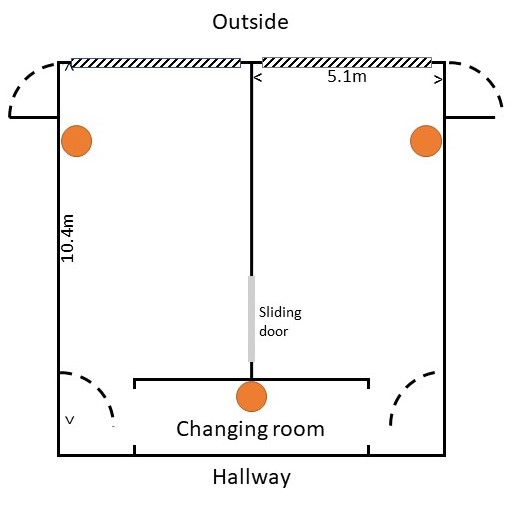

***Supplementary Figure 4.* Layout of two nursery sampling locations (groups 2 and 3) and placement of portable air cleaners*.*** The two main rooms were connected directly - through a sliding door which was most often open - and indirectly through the changing room*. In both groups, one air cleaner* was placed *on the floor and two at 1m above the floor (orange). The third nursery sampling location had a shorter room length of 9.6m and equal with. This resulted in a volume of 150m3, as opposed to 157m3 in groups 2 and 3.*

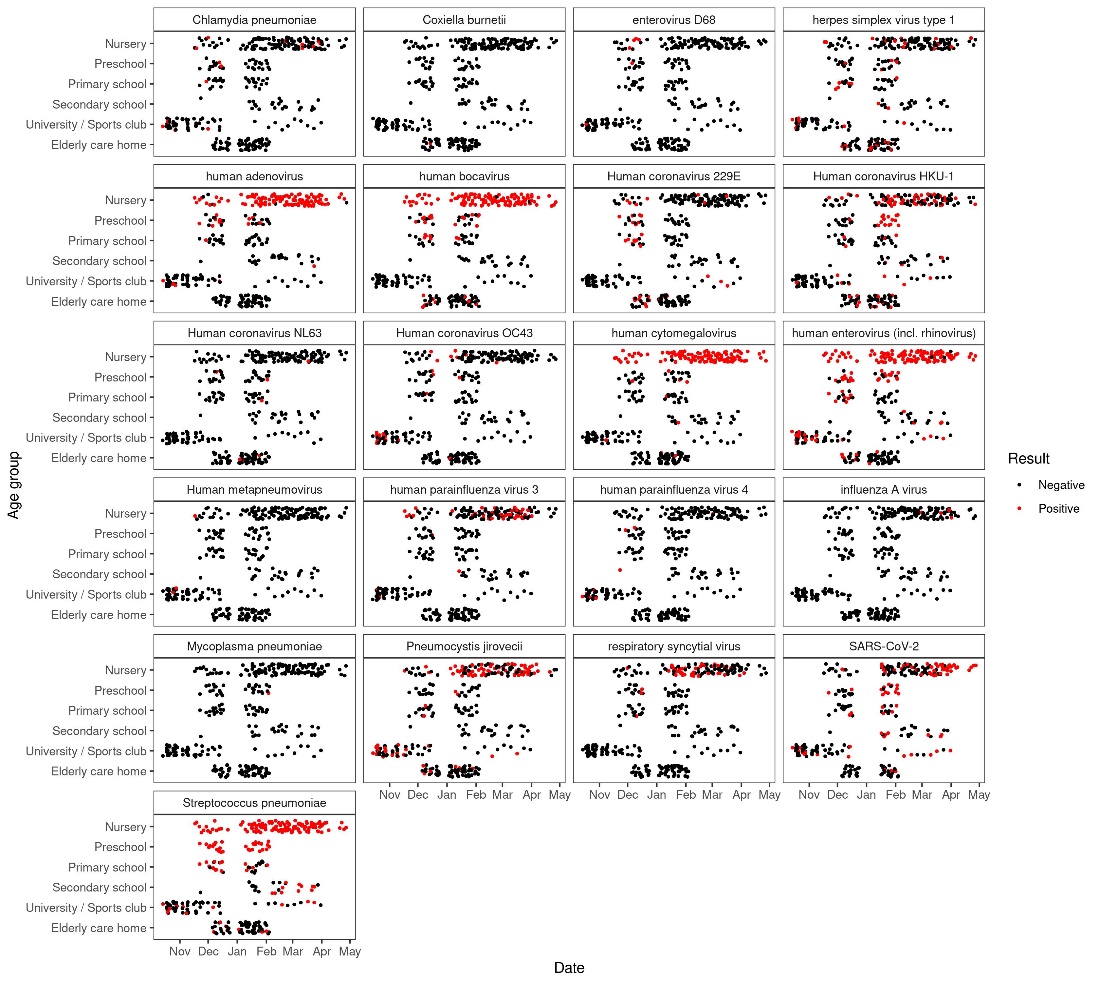

***Supplementary Figure 5. Overview of air samples and qPCR results for 29 pathogens.*** *This figure shows the positive (red) and negative (black) qPCR tests for 29 pathogens on environmental air samples. For SARS-CoV-2, it only shows the TaqPath qPCR results. Sample stratification (y axis) is by predominant age group: nursery (0-3y), preschool (3-6y), primary school (6-12y), secondary school (12-18y), adults (18-65y) and nursing homes (65+). The age groups 18y-25y and 25-65y were pooled as the latter represented only a limited number of samples. The x axis shows the timing of sampling. In October, sampling only took place in areas populated by university students. This expanded to a nursery, preschool, primary and secondary school in November. In December and January, we added several elderly care homes and one bar of a sports club. Sampling in university locations was reduced from the second part of December onwards due to exams. It pauzed around the Christmas holidays in schools. Sampling intensified in the nursery environment from January onwards as air cleaning was evaluated there.*

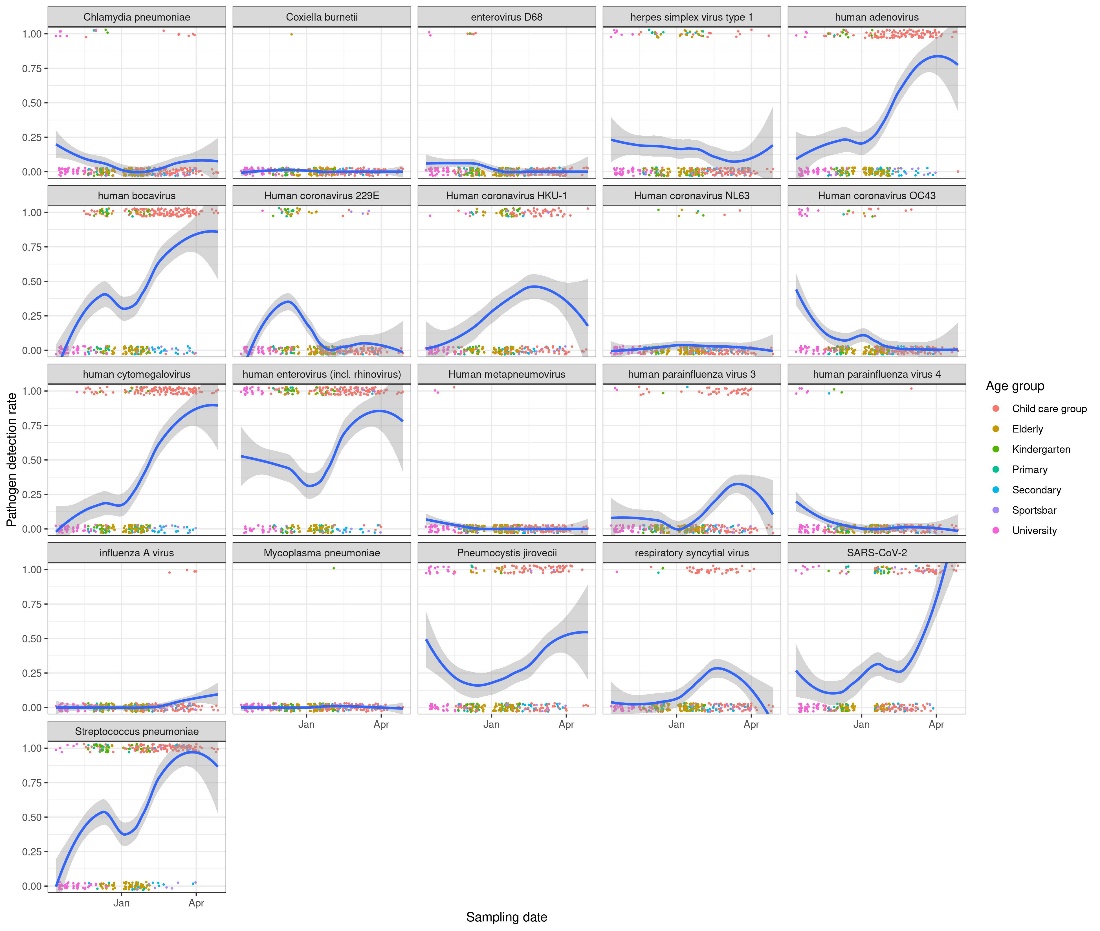

***Supplementary Figure*** ***6****.* ***LOcally weEighted Scatterplot Smoothing (LOESS) regression of the positivity rates of respiratory pathogens in all sampling sites.*** *Pathogens which were detected at least once are shown. Detection rates are likely influenced by the sites being sampled in the different time periods, besides epidemiology (Supplementary Figure 5).*

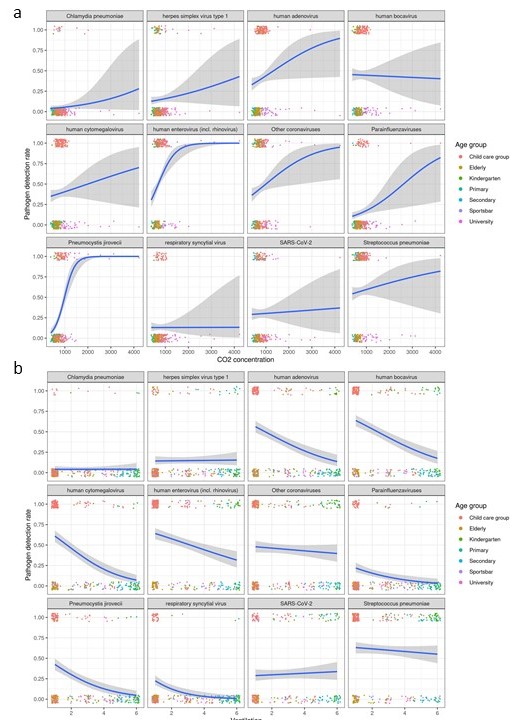

***Supplementary Figure 7. Univariate associations between mean*** *CO_2_* ***or natural ventilation and the detection of specific pathogens in indoor ambient air.*** *Panel a) shows the univariate association between the mean CO_2_ concentration and the probability of detecting a particular pathogen. Panel b) shows the univariate association between natural ventilation (as per the Linkert scale) and the probability of detecting a pathogen. In most cases, the association is positive between CO_2_ concentration and the risk of pathogen detection. The association is negative in most instances for natural ventilation. Counterintuitively, we don’t see this expected association between natural ventilation and detection for COVID-19, even though confidence intervals are quite small across the range. This may indicate the presence of a strong confounder, such as stricter compliance with non-pharmaceutical interventions when incidence was high. Indeed, the association is not significant in a multivariate analysis (Supplementary Table 6).*

### Supplementary tables

| **Site** | **Predominant age** | **Room Volume (m^3^)** | **HVAC present** | **Portable air cleaner present** | **Aimed sampling duration** | **Aimed weekly frequency** | **Sampling period** | **Total N samples** | **Positivity rate sample** | **Mean N of pathogens respiratory panel** |
| --- | --- | --- | --- | --- | --- | --- | --- | --- | --- | --- |
| Nursery location 1 | 0y-3y | 150,0 | NO | NO | 2h | 3 | 15/11/21- 27/4/22 | 60 | 98.93 | 7.00 |
| Nursery location 2 | 0y-3y | 157,5 | NO | YES | 2h | 3 | 17/01/22- 1/4/22 | 39 | 100.00 | 7.23 |
| Nursery location 3 | 0y-3y | 157,5 | NO | YES | 2h | 3 | 07/2/22- 1/4/22 | 24 | 100.00 | 7.75 |
| Preschool cafeteria | 3y-6y | 372,0 | NO | NO | 2h | 4 | 22/11/21- 4/2/22 | 26 | 100.00 | 4.50 |
| Preschool classroom | 3y-6y | 210,0 | NO | NO | 2h | 1 | 08/12/21- 2/2/22 | 4 | 100.00 | 4.75 |
| Primary school cafeteria | 6y-12y | 930,0 | NO | NO | 1-2h | 4 | 22/11/21- 1/2/22 | 25 | 68.00 | 1.92 |
| Primary school classroom | 6y-12y | 224,0 | NO | NO | 1h | 1 | 08/12/21- 2/2/22 | 4 | 100.00 | 4.75 |
| Secondary school cafeteria | 12y-18y | 912,0 | NO | NO | 2h | 4 | 23/11/21- 28/3/22 | 24 | 79.17 | 1.29 |
| University bar 1 | 18y-25y | 200,0 | NO | NO | 2h | 3 | 14/10/21 - 27/10/21 | 7 | 10.00 | 3.43 |
| University bar 2 | 18y-25y | 315,0 | NO | NO | 2h | 3 | 18/10/21- 10/11/21 | 7 | 100.00 | 3.29 |
| University bar 3 | 18y-25y | 198,0 | YES | NO | 2h | 3 | 18/10/21-10/11/21 | 10 | 80.00 | 2.60 |
| University cafeteria | 18y-25y | 7614,0 | YES | NO | 2h | 4 | 18/10/21-29/10/21 | 10 | 80.00 | 1.60 |
| University auditorium 1 | 18y-25y | 324,0 | NO | NO | 2h | 2 | 1/12/21-8/12/21 | 3 | 100.00 | 1.00 |
| University auditorium 2 | 18y-25y | 324,0 | NO | NO | 2h | 2 | 4/11/21-25/11/21 | 13 | 38.46 | 0.46 |
| University auditorium 3 | 18y-25y | 324,0 | NO | NO | 2h | 2 | 8/11/21-13/12/21 | 6 | 83.33 | 1.83 |
| University auditorium 4 | 18y-25y | 731,25 | NO | NO | 2h | 1 | 14/12/21-14/12/21 | 1 | 100.00 | 1.00 |
| Sports club bar | 25-65y | 324,0 | NO | NO | 2h | 1 | 20/01/22- 31/03/2022 | 10 | 90.00 | 2.00 |
| Elderly care home 1 | +65y | 273,0 | YES | NO | 2h | 4 | 9/12/21- 1/02/22 | 19 | 78.95 | 1.32 |
| Elderly care home 2 | +65y | 627,75 | YES | NO | 2h | 4 | 6/12/21- 4/02/22 | 35 | 77.14 | 1.71 |
| Elderly care home 3a | +65y | 600,0 | YES | NO | 2h | 2 | 06/1/22- 31/1/22 | 8 | 25.00 | 0.25 |
| Elderly care home 3b | +65y | 600,0 | YES | NO | 2h | 2 | 06/1/22 - 24/1/22 | 6 | 33.33 | 0.33 |

***Supplementary Table 1. The characteristics of the different sampling sites.*** *This table lists the different sampling sites. It shows their predominant age categories, room volume, the presence of heating, ventilation and air conditioning (HVAC), whether portable air cleaner were installed. It also lists the aimed sampling duration (accommodating site-specific schedules), the sampling period, the aimed sampling frequency and the total number of samples taken. The last columns list the crude positivity rate for any pathogen of all samples taken at a particular site and the average number of pathogens per sample. The aimed sampling time was 2 hours, unless the site-specific schedule dictated otherwise (e.g. lunch time in schools). The following sites were in the same institution: nursery groups 1 to 3; pre-school cafeteria, pre-school classroom and primary school cafeteria; elderly care home 3a and 3b. We focused on children and the elderly because of high incidence and morbidity from respiratory infections^24–26^. For university auditoria, rooms where high CO_2_ values were registered in the weeks prior to the start of the study were selected for inclusion. The nursery was adjacent to the University Hospital and hosts children of employees in the site.*

| **Site** | **HVAC type** | **Primary outdoor air fraction** | **Air transfer between zones** | **Recirculation within zone** | **Filter type (MERV)** | **Maximum volume displacement (m^3^/h)** | **Draining volume (m^3^)** | **Air changes per hour** | **On demand / continuous operation** |
| --- | --- | --- | --- | --- | --- | --- | --- | --- | --- |
| Elderly care home 1 | Stork-VDA200/4EC+WS-ventilator | NA | No | NA | NA | 1000 | 273 | 3.7 | On demand |
| Elderly care home 2 | TRANE CCEC 9/4.5-9/4.5 | ~ CO_2_ (ppm):  0% if < 400 60% if 400-800  100% if >1200 | No | Yes | 8-9 | 45000 | Unknown | Unknown | Continuous |
| Elderly care home 3a | CAIRplus 096.064IVBV | 100% | No | No | 13-14 | Unknown | 425 | Unknown | Continuous |
| Elderly care home 3b | CAIRplus 096.064IVBV | 100% | No | No | 13-14 | Unknown | 425 | Unknown | Continuous |
| University bar 3 | Airoxy arok 2500mm fan | 100% | NA | NA | NA | 1250 | 198 | 6.3 | On demand |
| University cafeteria | CAIRplus 160.160IVBV | 100% | No | NA | 13-14 | Unknown | 7493 | Unknown | ~ CO_2_ (ppm):  Flow increases when >800 ppm |
| Childcare group 2 | Three Blue PURE 221 (Blueair®) air cleaners | 0% | NA | 100% | 17 | 1,770 | 165 | 10.7 | Continuous |
| Childcare group 3 | Three Philips 3000i (Philips®) air cleaner | 0% | NA | 100% | 17 | 999 | 165 | 6.1 | Continuous |

***Supplementary table 2. HVAC/portable air cleaner characteristics.*** *The table shows the characteristics of heating, ventilation and air conditioning systems for sites with such a system. Filter maintenance followed the manufacturer's prescription. For the two nursery groups equipped with portable air cleaners, the site and system specifics are shown. Air changes per hour are theoretical, as they are computed using the maximum volume displacement and the volume of the drained area.*

| **Starting variables** | **Type** | **Missing data** | **Missing data handling** |
| --- | --- | --- | --- |
| Pathogen (grouped as per methods description) | factor | 0 |  |
| Predominant age group | factor | 0 |  |
| Month of sampling | factor | 0 |  |
| Sampling duration | numeric | 0 |  |
| Mean CO_2_ | numeric | 1/339 | omit observation |
| Relative humidity | numeric | 2/339 | omit observations |
| Temperature | numeric | 15/339 | impute missing data |
| Natural ventilation | numeric | 24/339 | impute missing data |
| Air cleaning | true/false | 0 | assume FALSE when missing |
| Mechanical ventilation | true/false | 0 |  |
| Mean number of attendees | numeric | 0 |  |
| Attendee density | numeric | 0 |  |
| Mask wearing | numeric | 64/339 | impute missing data |
| Vocalization | numeric | 73/339 | impute missing data |
| Weekly COVID-19 incidence Leuven | numeric | 0 |  |

***Supplementary table 3. Missing host, behavioral and environmental values and measures taken in their presence.*** *A total of 2/339 observations were omitted due to missing data, and 0/337 because the data could not be imputed following the method described in Supplementary Methods.*

| **Pathogen** | **N positive** | **N possibly non-specific** | **N retested** | **N retested positive** | **Decision possible non-specific** |
| --- | --- | --- | --- | --- | --- |
| influenza A virus | 4 | 0 | 0 | NA | NA |
| influenza B virus | 0 | 0 | 0 | NA | NA |
| respiratory syncytial virus A & B | 43 | 0 | 0 | NA | NA |
| *human metapneumovirus* | 3 | 0 | 0 | NA | NA |
| human parainfluenza virus 1 | 0 | 0 | 0 | NA | NA |
| human parainfluenza virus 2 | 3 | 3 | 3 | 0 | Classify as negative |
| human parainfluenza virus 3 | 43 | 0 | 0 | NA | NA |
| human parainfluenza virus 4 | 9 | 0 | 0 | NA | NA |
| human adenovirus | 134 | 9 | 5 | 3 | Classify as positive |
| human enterovirus (incl. rhinovirus) | 182 | 0 | 0 | NA | NA |
| human cytomegalovirus | 131 | 1 | 1 | 0 | Classify as negative |
| human parechovirus | 5 | 5 | 5 | 0 | Classify as negative |
| *Human coronavirus NL63* | 13 | 13 | 5 | 0 | Classify as negative |
| *Human coronavirus 229E* | 37 | 28 | 5 | 4 | Classify as positive |
| *Human coronavirus OC43* | 32 | 21 | 5 | 2 | Classify as negative |
| herpes simplex virus type 1 | 50 | 0 | 0 | NA | NA |
| herpes simplex virus type 2 | 0 | 0 | 0 | NA | NA |
| enterovirus D68 | 6 | 0 | 0 | NA | NA |
| SARS-CoV-2 | 10 | 2 | 0 | NA | NA |
| *Mycoplasma pneumoniae* | 1 | 0 | 0 | NA | NA |
| *Human coronavirus HKU1* | 98 | 85 | 5 | 5 | Classify as positive |
| *Pneumocystis jiroveci* | 100 | 0 | 0 | NA | NA |
| *Coxiella burnetii* | 4 | 3 | 3 | 1 | Classify as negative |
| MERS | 3 | 3 | 3 | 0 | Classify as negative |
| *Chlamydia pneumoniae* | 23 | 23 | 5 | 1 | Classify as negative |
| *Chlamydia psittaci* | 1 | 1 | 1 | 0 | Classify as negative |
| *Streptococcus pneumoniae* | 195 | 0 | 0 | NA | NA |
| *Legionella pneumophila* | 0 | 0 | 0 | NA | NA |
| human bocavirus | 152 | 29 | 5 | 4 | Classify as positive |

***Supplementary table 4.*** *Results of respiratory panels with CT values compatible with possible non-specific amplification. CT values compatible with possible non-specific amplification according to clinical cutoffs for respiratory samples, and the results of repeated testing using qPCR with fresh primer probe mix. A total of 226 CT values were possibly compatible with non-specific amplification. After the initial rerun of up to 5 samples with the lowest CT values per pathogen (less than 5 samples were run per pathogen only when less than 5 samples had possible non-specific amplification for that pathogen). If at least 60% of samples returned positive for, we retained the initial result. If less than 60% of samples returned positive, all samples with possible non-specific amplification for that pathogen were reclassified as negative.*

| a) **Pathogen detection (all pathogens)** | | | |
| --- | --- | --- | --- |
| **Remaining variables** | **p-value** | **Effect size (odd ratio and 95% CI)** | **Direction** |
| **LRM** | | | |
| Pathogen | <0.0001 |  |  |
| Age group | <0.0001 |  |  |
| Month | 0.0025 |  |  |
| CO_2_ | 0.0015 | 1.09 (CI 1.03-1.15) per 100 increase in CO_2_ | positive |
| Natural ventilation | 0.0097 | 0.89 (CI 0.80-0.97) per step increase | negative |
| **MLRM (random effect: sample ID)** | | | |
| Pathogen | <0.0001 |  |  |
| Age group | <0.0001 |  |  |
| Month | 0.0025 |  |  |
| CO_2_ | 0.0015 | 1.09 (CI 1.03-1.15) per 100 increase in CO_2_ | positive |
| Natural ventilation | 0.0097 | 0.88 (CI 0.80-0.97) per step increase | negative |
| **GEE (grouped by sample ID)** | | | |
| Pathogen | <0.0001 |  |  |
| Age group | <0.0001 |  |  |
| Month | 0.0016 |  |  |
| CO_2_ | 0.0004 | 1.09 (CI 1.04-1.14) per 100 increase in CO_2_ | positive |
| Natural ventilation | 0.0044 | 0.88 (CI 0.80-0.96) per step increase | negative |
| b) **Pathogen concentration (qPCR CT of positive samples, all pathogens)** | | | |
| **Remaining variables** | **p-value** | **Effect size (change in CT value and 95% CI)** | **Direction** |
| **LRM** | | | |
| Pathogen | <0.0001 |  |  |
| Age group | <0.0001 |  |  |
| Month | 0.002 |  |  |
| CO_2_ | <0.0001 | -0.08 (CI -0.12 to -0.04) per increase of 100 | negative |
| Air cleaning | 0.00054 | 0.58 (CI 0.25-0.91) | positive |
| **MLRM (random effect: sample ID)** | | | |
| Pathogen | <0.0001 |  |  |
| Age group | <0.0001 |  |  |
| Month | 0.0067 |  |  |
| CO_2_ | 0.0003 | 0.08 (CI -0.12 to -0.04) per increase of 100 in CO_2_ | negative |
| Air cleaning | 0.002 | 0.58 (CI 0.21-0.95) | positive |

***Supplementary table 5. Impact of environmental parameters on bioaerosol load in a logistic regression model (LRM), mixed logistic regression model (MLRM) and Generalized Estimating Equations model (GEE).*** *The significance levels and effect sizes of the input variables are very similar across models (see also Table 1).*

| **Pathogen** | **Variable** | **Effect estimate** | **p-value** | **Odds ratio (95% CI)** |
| --- | --- | --- | --- | --- |
| human adenovirus | NA | NA | NA | NA |
| human bocavirus | Mean CO_2_ | -0.559 | 0.017717 | 0.572 (0.36 - 0.908) |
| *Chlamydophila pneumoniae* | NA | NA | NA | NA |
| human cytomegalovirus | NA | NA | NA | NA |
| human enterovirus (incl. rhinovirus) | Mean CO_2_ | 0.131 | 0.027575 | 1.14 (1.015 - 1.281) |
| herpes simplex virus type 1 | NA | NA | NA | NA |
| Other coronaviruses | Mean CO_2_ | 0.148 | 0.006872 | 1.159 (1.042 - 1.291) |
| human parainfluenzavirus 1 | NA | NA | NA | NA |
| *Pneumocystis jiroveci* | Mean CO_2_ | 0.447 | 0.000033 | 1.564 (1.266 - 1.932) |
| *Pneumocystis jiroveci* | Mean natural ventilation | -0.393 | 0.031706 | 0.675 (0.472 - 0.966) |
| respiratory syncytial virus | Mean natural ventilation | -0.885 | 0.007045 | 0.413 (0.217 - 0.786) |
| *Streptococcus pneumoniae* | Mean CO_2_ | 0.105 | 0.031713 | 1.111 (1.009 - 1.222) |
| SARS-CoV-2 | NA | NA | NA | NA |

***Supplementary table 6 shows the influence of environmental factors on individual pathogen detection in a generalized linear model.*** *A generalized linear model was built for each specific pathogen. Covariates were those independently associated with pathogen presence in previous models. We repeated backward elimination while always retaining the sampling Month and dominant age group. The table lists the pathogen of interest and the retained variables after backward elimination. The last column shows the odds of detection per 100 ppm increase in CO_2_ or stepwise increase in natural ventilation and the corresponding 95% CI.*

| **Pathogen** | **Retained variable** | **p-value** | **Effect estimate (95% CI)** |
| --- | --- | --- | --- |
| human adenovirus | Mean CO_2_ | 1.58E-02 | -0.33(-0.5936 - -0.0658) |
| human bocavirus | Mean CO_2_ | 6.37E-05 | -0.437(-0.6444 - -0.2291) |
| human bocavirus | Air cleaning | 4.68E-02 | 0.7(0.0159 - 1.3845) |
| *Chlamydophila pneumoniae* | NA | NA | NA |
| human cytomegalovirus | Mean CO_2_ | 3.12E-02 | -0.21(-0.3984 - -0.0212) |
| human cytomegalovirus | Air cleaning | 1.88E-03 | 0.964(0.37 - 1.5587) |
| human enterovirus (incl. rhinovirus) | NA | NA | NA |
| herpes simplex virus type 1 | NA | NA | NA |
| Other coronaviruses | Air cleaning | 1.94E-03 | 2.041(0.7754 - 3.3075) |
| Parainfluenza viruses | NA | NA | NA |
| *Pneumocystis jiroveci* | NA | NA | NA |
| *respiratory syncytial virus* | Mean CO_2_ | 3.55E-02 | 0.474(0.0493 - 0.899) |
| *Streptococcus pneumoniae* | Mean CO_2_ | 3.04E-03 | -0.122(-0.2023 - -0.0426) |
| *Streptococcus pneumoniae* | Air cleaning | 2.26E-05 | 1.106(0.6082 - 1.6047) |
| SARS-CoV-2 | NA | NA | NA |

***Supplementary table 7 shows the influence of environmental factors on individual pathogen concentration (expressed as Ct value) in a linear regression model.*** *A linear regression model was built for each specific pathogen. Covariates were those independently associated with pathogen concentration in previous models. We repeated backward elimination while always retaining the sampling Month and dominant age group. The table lists the pathogen of interest and the retained variables after backward elimination. The last column shows the estimated change in CT value per 100 ppm increase in CO_2_ concentration or stepwise increase in natural ventilation, and the 95% CI.*

| **Location** | **Cleaning phase** | **Change in CT value (95% CI)** | **p-value** |
| --- | --- | --- | --- |
| Group 1 | Wednesdays-Mondays | 0.016  (-0.48 - 0.51) | 0.95 |
|  | Fridays-Mondays | -0.093  (-0.59 - 0.40) | 0.71 |
| Group 2 | Wednesdays-Mondays | 1.22  (0.65 - 1.79) | < 0.0001 |
|  | Fridays-Mondays | 1.34  (0.56 - 1.70) | 0.00018 |
| Group 3 | Wednesdays-Mondays | 0.33  (-0.32 - 0.98) | 0.31 |
|  | Fridays-Mondays | 1.02  (0.37 - 1.67) | 0.003 |

***Supplementary table 8. The influence of air cleaning on pathogen concentrations in an interventional comparison, assessed through a mixed-model linear regression.*** *Repeated samples in different cleaning phases were used as input. The change in mean CT value of all positive respiratory pathogens against the baseline value on Mondays was the model outcome.*

| Multiplex PCR | Pathogen | Primer/probe | Primer/probe sequence (5’ 🡪 3’) | Concentra-tion (nµ) | Target gene | Amplicon size (bp) | CT cut-off positive versus negative |
| --- | --- | --- | --- | --- | --- | --- | --- |
| 1 | influenza A virus | Forward primer | TCTCATGGAATGGCTAAAGACAAG | 300 | Matrix protein gene | 116 | NA |
|  |  | Reverse primer | CAAAGCGTCTACGCTGCAGT | 300 |  |  |  |
|  |  | Probe | FAM-TTCACGCTCACCGTGC-MGB | 200 |  |  |  |
|  | influenza B virus | Forward primer | AATACGGTGGATTAAACAAAAGCA | 300 | Hemagluttinin gene | 170 | NA |
|  |  | Reverse primer | ACCAGCAATAGCTCCGAAGAA | 300 |  |  |  |
|  |  | Probe | NED-TATATTTGGTTCCATTGGC-MGB | 200 |  |  |  |
|  | PDV  (RNA IC) | Forward primer | GGTGGGTGCCTTTTACAAGAAC (^6^) | 250 | Hemagluttinin gene | 83 | NA |
|  |  | Reverse primer | ATCTTCTTTCCTCAACCTCGTCC (^6^) | 250 |  |  |  |
|  |  | Probe | VIC-ATGCAAGGGCCAATT-MGB (^6^) | 100 |  |  |  |
| 2 | respiratory syncytial virus A +  RSV B | Forward primer | AATACAGCCAAATCTAACCAACTTTACA (^7^) | 300 | RNA polymerase gene | 94 | NA |
|  |  | Reverse primer | GCCAAGGAAGCATGCAATAAA (^7^) | 300 |  |  |  |
|  |  | Probe A | FAM-TGATGTGCTATTGTGCACTA-MGB (^7^) | 100 |  |  |  |
|  |  | Probe B | FAM-CACTATTCCTTACTAAAGATGTC-MGB (^7^) | 100 |  |  |  |
|  | HMPV | Forward primer | GCYGTYAGCTTCAGTCAATTCAA | 500 | Fusion protein gene | 75 | NA |
|  |  | Reverse primer | TGTTATYCCWGCATTGTCTGA | 500 |  |  |  |
|  |  | Probe | NED-CTAAATGTTGTGCGGCAAT-MGB | 200 |  |  |  |
| 3 | parainfluenza virus 1 | Forward primer | CCTTCATTATCAATTGGTGATGCA | 500 | Hemagglutinin-neuraminidase gene | 180 | NA |
|  |  | Reverse primer | CCTGTTGTCGTTGATGTCATAGGT | 500 |  |  |  |
|  |  | Probe | FAM-TCAAACTTAATCACTCAAGGAT-MGB | 100 |  |  |  |
|  | parainfluenza virus 2 | Forward primer | CATTTACCTAAGTGATGGAATCAATCG | 500 | Hemagglutinin-neuraminidase gene | 141 | 36.5 |
|  |  | Reverse primer | GCAAGTCTCAGTTCAGCTAGATCAGT | 500 |  |  |  |
|  |  | Probe | NED-AAAGCTGTTCAGTCACTG-MGB | 150 |  |  |  |
| 4 | parainfluenza virus 3 | Forward primer | CAAAGTTGATGAAAGATCAGATTATGC | 500 | Hemagglutinin-neuraminidase gene | 169 | NA |
|  |  | Reverse primer | GTAGTATATCCCTGGTCCAACAGATG | 500 |  |  |  |
|  |  | Probe | FAM-CAATCTCRACAACAAGATT-MGB | 100 |  |  |  |
|  | parainfluenza virus 4 | Forward primer | GGAGCAAAAGAYTCATACACAATAACTTACT | 500 | Hemagglutinin-neuraminidase gene | 116 | NA |
|  |  | Reverse primer | CTAGGAATTAARATTTCACAATCTTCAGAA | 500 |  |  |  |
|  |  | Probe | NED-AAATGCACTCTGTATAAGTC-MGB | 150 |  |  |  |

| Multiplex PCR | Pathogen | Primer/probe | Primer/probe sequence (5’ 🡪 3’) | Concentra-tion (nµ) | Target gene | Amplicon size (bp) | CT cut-off positive versus negative |
| --- | --- | --- | --- | --- | --- | --- | --- |
| 5 | human adenovirus | Forward primer | TYGARGTGGAYCCCATGGAYGAG | 800 | Hexon gene | 112 | 39.0 |
|  |  | Reverse primer | CGCAGGTAGACBGCYTCRATGA | 800 |  |  |  |
|  |  | Probe 1 | FAM-ACGTCGAAGACTTC-MGB | 100 |  |  |  |
|  |  | Probe 2 | FAM-ACGTCGAAAACTTC-MGB | 100 |  |  |  |
|  |  | Probe 3 | FAM-CACGTCAAAGACTTC-MGB | 100 |  |  |  |
|  | human enterovirus (inc. hino-virus) | Forward primer 1 | CAWGGTGYGAAGAGYCTATTGAGCT | 500 | Polyprotein gene | 150-154 | NA |
|  |  | Forward primer 2 | GTGTGAAGASCCSMGTGYGCT | 500 |  |  |  |
|  |  | Reverse primer | GAAACACGGACACCCAAAGTAGT | 500 |  |  |  |
|  |  | Probe | VIC-TCCGGCCCCTGAATGYGGCTAA-TAMRA | 200 |  |  |  |
| 6 | human cytomegalovirus | Forward primer | CGTAACGTGGACCTGACGTTT | 250 | Major capsid protein gene | 148 | 37.0 |
|  |  | Reverse primer | CACGGTCCCGGTTTAGCA | 250 |  |  |  |
|  |  | Probe | FAM-TATCTGCCCGAGGATCGCGGTTACA-TAMRA | 200 |  |  |  |
|  | Human parechovirus | Forward primer | CASWWGCCTCTGGGSCCAAAAG (^8^) | 500 | Polyprotein gene | 189 | 36.0 |
|  |  | Reverse primer | GGCCCCWGRTCAGATCCAYAGT (^8^) | 500 |  |  |  |
|  |  | Probe | Cy5-CCTRYGGGTACCTYCWGGGCATCCTTC-BHQ3 (^8^) | 200 |  |  |  |
|  | PhHV-1  (DNA IC) | Forward primer | GGGCGAATCACAGATTGAATC (^9^) | 100 | Glycoprotein B gene | 89 | NA |
|  |  | Reverse primer | GCGGTTCCAAACGTACCAA (^9^) | 100 |  |  |  |
|  |  | Probe | VIC-TCCGCCACCATCTG-MGB | 100 |  |  |  |
| 7 | *Human coronavirus NL63* | Forward primer | CAGGGCTGACAAGCCTTCTCA (^10^) | 500 | Nucleocapsid protein gene | 144 | 35.0 |
|  |  | Reverse primer | GCATCAACACCATTCTGAACAAGA (^10^) | 500 |  |  |  |
|  |  | Probe | FAM-CGTTGGAAGCGTGTTCCTACCAGAGAGG-BHQ-1 (^10^) | 200 |  |  |  |
|  | *Human coronavirus 229E* | Forward primer | TGGAAGTGCAGGTGTTGTGGC (^10^) | 500 | Nucleocapsid protein gene | 99 | 34.5 |
|  |  | Reverse primer | TGACTATCAAACAGCATAGCAGCTG (^10^) | 500 |  |  |  |
|  |  | Probe | Cy5-CCACAATTTGCTGAGCTTGTGCCGTC-BHQ-3 (^10^) | 200 |  |  |  |
|  | *Human coronavirus OC43* | Forward primer | CGATGAGGCTATTCCGACTAGGT (^11^) | 500 | Nucleocapsid protein gene | 75 | 35.5 |
|  |  | Reverse primer | CCTTCCTGAGCCTTCAATATAGTAACC (^11^) | 500 |  |  |  |
|  |  | Probe | VIC-TCCGCCTGGCACGGTACTCCCT-TAMRA (^11^) | 200 |  |  |  |

| Multiplex PCR | Pathogen | Primer/probe | Primer/probe sequence (5’ 🡪 3’) | Concentra-tion (nµ) | Target gene | Amplicon size (bp) | CT cut-off positive versus negative |
| --- | --- | --- | --- | --- | --- | --- | --- |
| 8 | herpes simplex virus 1 & 2 | Forward primer | AACCTGGGRTTCCTGATGCA | 250 | Nonfunctional glycoprotein D gene (US6) | 84 |  |
|  |  | Reverse primer | CTCCGTCCAGTCGTTTATCTTCAC | 250 |  |  |  |
|  |  | Probe HSV-1 | FAM-TTTGAGACCGCCGGCACGTAC-BHQ-1 | 100 |  |  | NA |
|  |  | Probe HSV-2 | Cy5-CCTTCGAGACCGCGGGTACGTA-BHQ-3 | 100 |  |  | 35.0 |
|  | enterovirus D68 | Forward primer | TGGCGGCCTACTCATGG (^12^) | 500 | Polyprotein gene | 64 | NA |
|  |  | Reverse primer | AATAGACTCTTCACACCTTGTTCATGT (^12^) | 500 |  |  |  |
|  |  | Probe | NED-AAAACCATGAGACGCT-MGB (^12^) | 200 |  |  |  |
|  | SARS-CoV-1 + SARS-CoV-2 | Forward primer | CWGGCATACCWAAGGACATGACCTA | 500 | ORF1ab polyprotein gene | 147 | NA |
|  |  | Reverse primer | CKACATCRAAGCCAATCCA | 500 |  |  |  |
|  |  | Probe | VIC-TTTATCACCCGCGAAGAA-MGB | 200 |  |  |  |
| 9 | *Mycoplasma pneumoniae* | Forward primer | AGGCTTCAAGTGGACAAAGTGAC | 250 | Adhesin P1 gene | 76 | NA |
|  |  | Reverse primer | GATTGTYCCTGCTGGYCCAT | 250 |  |  |  |
|  |  | Probe | FAM-ACCACACCAAGTTCA-MGB | 200 |  |  |  |
|  | *Human coronavirus HKU1* | Forward primer | CACTTCTATTCCCTCCGATGTTTC (^10^) | 500 | Nucleocapsid protein gene | 129 | 32.5 |
|  |  | Reverse primer | TTAGAAGCAGACCTTCCTGAGCC (^10^) | 500 |  |  |  |
|  |  | Probe | Cy5-CGCCTGGTACGATTTTGCCTCAAGGCT-BHQ-3 (^10^) | 200 |  |  |  |
| 10 | *Pneumocystis jiroveci* | Forward primer | GCACTGAATATCTCGAGGGAGTATG | 250 | Large subunit ribosomal RNA gene | 145 | NA |
|  |  | Reverse primer | TTGGGAGCTTTAATTACTGTTCTGG | 250 |  |  |  |
|  |  | Probe | FAM-TGTTTCCCTTTCGACTATC-MGB | 200 |  |  |  |
|  | *Coxiella burnetii* | Forward primer | CGATAGCCCGATAAGCATCAAC (^13^) | 250 | IS1111 transposase gene | 88 | 36.7 |
|  |  | Reverse primer | TGCATTCGTATATCCGGCATC (^13^) | 250 |  |  |  |
|  |  | Probe | NED-TCATCAAGGCACCAATG-MGB (^13^) | 200 |  |  |  |
|  | MERS | Forward primer | GCAACGCGCGATTCAGTT (^14^) | 500 | Envelope gene (upE) | 92 | 35.5 |
|  |  | Reverse primer | GCCTCTACACGGGACCCATA (^14^) | 500 |  |  |  |
|  |  | Probe | Cy5-CTCTTCACATAATCGCCCCGAGCTCG-BHQ-3 (^14^) | 200 |  |  |  |

| Multiplex PCR | Pathogen | Primer/probe | Primer/probe sequence (5’ 🡪 3’) | Concentra-tion (nµ) | Target gene | Amplicon size (bp) | CT cut-off positive versus negative |
| --- | --- | --- | --- | --- | --- | --- | --- |
| 11 | *Chlamydia pneumoniae* | Forward primer | CATCCGTGTCGGAGCTAACGT (^15^) | 250 | 16S rRNA gene | 184 | 34.5 |
|  |  | Reverse primer | TGCGGAAAGCTGTATTTCTACAGTT (^15^) | 250 |  |  |  |
|  |  | Probe | FAM-ATGCCGCCTGAGGAGTACACTCGC-BHQ1 (^15^) | 200 |  |  |  |
|  | *Chlamydia psittaci* | Forward primer | GCCATCATGCTTGTTTCGTTT (^16^) | 250 | Inclusion membrane protein A gene | 74 | 40.0 |
|  |  | Reverse primer | CGGCGTGCCACTTGAGA (^16^) | 250 |  |  |  |
|  |  | Probe | VIC-TTGTCATTATGGTGATTCAGGA-MGB (^16^) | 200 |  |  |  |
|  | *Streptococcus pneumoniae* | Forward primer | ACGCAATCTAGCAGATGAAGCA (^17^) | 250 | Autolysin gene (lytA) | 75 | 36.0 |
|  |  | Reverse primer | TCGTGCGTTTTAATTCCAGCT (^17^) | 250 |  |  |  |
|  |  | Probe | Cy5-TGCCGAAAACGCTTGATACAGGGAG-BHQ3 (^17^) | 200 |  |  |  |
| 12 | *Legionella pneumophila* | Forward primer | TCCGGAAGCAATGGCTAAAG | 250 | Macrophage infectivity potentiator gene | 69 | 35.5 |
|  |  | Reverse primer | TGCTGTTCGGTTAAAGCCAAT | 250 |  |  |  |
|  |  | Probe | FAM-CATGCAAGACGCTATGAGTGGCGCT-TAMRA | 200 |  |  |  |
|  | human bocavirus | Forward primer | GCACAGCCACGTGACGAA (^18^) | 250 | Nonstructural protein 1 gene | 76 | 34.5 |
|  |  | Reverse primer | TGGACTCCCTTTTCTTTTGTAGGA (^18^) | 250 |  |  |  |
|  |  | Probe | Cy5-TGAGCTCAGGGAATATGAAAGACAAGCATCG-BHQ-3 (^18^) | 200 |  |  |  |

***Supplementary table 9. List of primer and probe sequences used in the 12 multiplex PCRs for the respiratory panel.*** *Ambiguity codes: R: A or G; Y: C or T; W: A or T; S: C or G; K: G or T. PDV: phocine distemper virus; IC: internal control; PhHV: phocine herpesvirus. Cut-off CT values are based on the reexamination of samples with possible non-specific amplification and the results from those reruns. NA signifies no possible non-specific amplification has taken place so that no cutoff CT has had to be determined.*

4.	Indoor air quality probe for CO2, temperature, humidity and absolute pressure | Testo international. https://www.testo.com/en/iaq-probe-to-assess-indoor-air-quality-co2-humi/p/0632-1535.

5.	Elektroniksystem i Umeå AB.
